# Perspectives of Osteopathic Medical Students on Preclinical Urology Exposure: A Single Institution Cross Sectional Survey

**DOI:** 10.1101/2024.02.03.24302283

**Authors:** Ryan Wong, Harvey N. Mayrovitz

## Abstract

**Context:** There is an increasing number of medical school graduates opting for surgical specialties and the osteopathic applicant match rate for urology is lower than that of allopathic applicants. Factors influencing this may include a lack of interest, perceived challenges in matching into urology, insufficient urology mentorship, limited research opportunities, and inadequate osteopathic representation in urology.

**Objective:** To assess osteopathic medical students’ perspectives on pursuing urology and enhancing preclinical exposure to and knowledge of urology.

**Methods:** A 20-question survey addressing experiences and the factors influencing osteopathic medical students’ specialty selection and their interest in and perception of urology was designed by the investigators on Research Electronic Data Capture software. This survey was distributed via email listserv to all current osteopathic medical students attending Nova Southeastern University Dr. Kiran C. Patel College of Osteopathic Medicine over two months. Responses were collected and analyzed using Fisher’s exact test.

**Results:** Of 150 respondents, 91% found mentors crucial in selecting a medical specialty, 95% emphasized the importance of early exposure, and 68% lacked familiarity with urology, with more M1 students unfamiliar compared to M2 (70.37% vs. 59.02%). A larger proportion of combined M1 and M2 (preclinical) students are considering urology as a specialty compared to M3 and M4 (clinical) students who are actively on rotations (56.52% vs. 28.57%; *p* = 0.0064). Also, a greater percentage of males are considering urology compared to females (64.15% vs. 42.71%; *p* = 0.0164). Among those considering urology (n = 75), 57.3% lack awareness of urology’s scope, and 84% report no preclinical discussions with urologists. Those students who report they are considering urology value early exposure significantly more than others (98.67% vs. 78.67%; *p* = 0.0001). They also express greater interest in having a core urology course (73.33% vs. 38.67%; *p* < 0.0001). More urology considering students are interested in extracurricular urology-related workshops, seminars, or conferences (61.33% vs. 17.33%; *p* <0.0001). Students who are considering urology as a specialty show greater interest in having a mentorship program (85.33% vs. 28%; *p* < 0.0001).

**Conclusion:** Results suggested that increased urology exposure during the preclinical years is important. Urology elective offerings and urology mentorship are of high interest among those considering urology. However, additional investigation is needed to determine the impact of preclinical urology curricula implementation on urology match outcomes.

## INTRODUCTION

Urology stands as an independent residency program in the United States, open to medical students who have completed either allopathic or osteopathic medical school. It is widely regarded as a highly competitive surgical sub-specialty.^1^ Urology residency programs engage in a unique early match process, distinct from the National Resident Matching Program (NRMP), facilitated through the American Urological Association (AUA). In 2020, the American Association of Colleges of Osteopathic Medicine (AACOM) and the American Osteopathic Association (AOA) merged, resulting in the establishment of a unified graduate medical education accreditation system.^2^ According to data provided by the AUA, the match rate for urology in 2023 was 75.39%, offering a total of 383 available residency positions.^3^ Prior to the 2024 urology match, the AUA did not report on the distinction between allopathic and osteopathic medical students within the match data.

Before the merger of the match system, there were 11 programs exclusively available to osteopathic medical students.^4^ In contrast to allopathic medical schools, many osteopathic institutions lack urology residency programs within their campuses.^4^ Consequently, this situation underscores the obstacles faced by aspiring urology trainees who seek proximity to their academic institutions for educational opportunities. Moreover, it has been reported that an enduring bias against osteopathic medical students has persisted over time, largely influenced by the historical prevalence of osteopathic physicians predominantly entering primary care fields.^5^ This bias has occasionally led to misconceptions regarding the qualifications of osteopathic graduates, with their credentials not always being viewed as equivalent to those of their allopathic counterparts. A recent study has shed light on this issue by revealing that there is no discernible difference in the outcomes of orthopedic in-training exam scores between DO and MD degree-holding residents.^6^

While there is a growing number of medical school graduates opting for surgical specialties today and low historical match rates for osteopathic medical students in competitive specialties, the osteopathic medical student match rate in urology is also lower than allopathic applicants.^2,7^ Possible contributing factors may include a lack of interest, perceived difficulty in securing a urology residency, insufficient support for osteopathic medical students in this field, limited research opportunities, or inadequate exposure to the full scope of what urology entails.^4,8^ Studies have underscored the significance of the preclinical years in medical school as a crucial phase for exposing students to various specialties.^9,10^

To the best of our knowledge, there is no study examining osteopathic medical students’ attitudes and perspectives on preclinical exposure to urology. The objective of our present study is to determine whether a lack of preclinical exposure to and knowledge of urology impacts osteopathic medical student’s decision to pursue urology as a specialty based on a single institutional analysis.

## METHODS

This study was approved and deemed exempt by Nova Southeastern University’s Institutional Review Board (NSU IRB Study: 2023-391). A survey-based methodology was utilized for this study. Twenty multiple choice questions were created on Research Electronic Data Capture (REDCap), a secure web-based software, containing categorical variables in the answer choices as shown in Supplementary Table S1. Survey questions were designed to assess osteopathic medical students’ interest in and perception of urology and preclinical exposure. The first two questions asked for the respondent’s medical school year and gender. The next four questions pertained to the respondent’s perspectives regarding overall medical specialty decisions. The final 14 questions gauged the respondent’s attitudes and perspectives on urology interest and preclinical exposure.

The survey was disseminated through university email lists to all current first, second, third, and fourth year osteopathic medical students enrolled at Nova Southeastern University Dr. Kiran C. Patel College of Osteopathic Medicine. Students at both campuses (Davie and Tampa) were included. Responses were voluntary and anonymous. Over the course of two months, five emails were sent. Responses that respondents completed that took them more than one minute and also had all questions answered were included in the final analysis. If the respondent answered the questions in less than one minute these responses were excluded from analysis to mitigate against responses that did not have time for thoughtful consideration.

### Statistical Analyses

Descriptive statistics were employed to analyze all of the data, while more detailed data quantification involved the use of Fisher’s exact test for categorical data. The analysis encompassed the comparison of specific groups: M1/M2 vs M3/M4 (preclinical vs clinical status), M1 vs M2, and Male vs Female. Additionally, the distinction between those considering urology and not considering urology was derived from responses to the question: “Are you currently considering urology as a potential specialty choice?” Those who responded with “Yes, I am seriously considering urology as a specialty” or “I am open to considering urology but exploring other options as well” were classified as considering urology.

To enhance data interpretation, scaled survey responses were transformed into binary categories for analysis in this study. The specific survey questions and their corresponding binary outcomes are presented in Supplementary Table S1. This categorization facilitated the use of Fischer’s exact test to explore potential associations between the binary variables. GraphPad Prism® 10 Software was utilized for all statistical analyses. All *p* values < 0.05 were considered statistically significant.

## RESULTS

A total of 152 responses were received. One was excluded due to an incomplete response, and another one was excluded due to a survey completion time of less than one minute. 150 responses were included in the study for analysis. 53 (35.3%) respondents identified as male, 96 (64%) respondents identified as female, and 1 (0.7%) respondent identified as other (Table 1).

**Table 1:** Characteristics of survey respondents.

| *n* = 150 |  |  |
| --- | --- | --- |
| Gender, No. (%) | Male | 53 (35.3) |
|  | Female | 96 (64) |
|  | Other | 1 (0.7) |
| Year in medical school, No. (%) | 1 | 54 (36) |
|  | 2 | 61 (40.67) |
|  | 3 | 19 (12.67) |
|  | 4 | 16 (10.67) |
| Most important factor when deciding a medical specialty, No. (%) | Personal interest and passion | 93 (62) |
|  | Lifestyle and work-life balance | 52 (34.67) |
|  | Job market and demand | 3 (2) |
|  | Potential earnings | 2 (1.33) |
|  | Influence of mentors | 0 (0) |

When looking at the distribution of respondents’ current year in medical school, 54 (36%) were M1, 61 (40.67%) were M2, 19 (12.67%) were M3, and 16 (10.67%) were M4 (Table 1). 62% of respondents indicated personal interest and passion as the most important factors when deciding a medical specialty while 34.67% of respondents indicated lifestyle and work-life balance as the most important factor (Table 1).

### Factors Influencing Specialty of Choice

A majority of respondents (91%) express the belief that mentors and advisors play a significant role in shaping their decision to pursue a specific medical specialty. Notably, a higher proportion of females (62%) perceive mentors as influential compared to males (30%) (*p*=0.0271). Regarding the importance of early exposure in the decision of pursuing a certain medical specialty, 95% of respondents affirm its significance. Gender-wise, a larger percentage of females (62%) emphasize the importance of early exposure compared to males (34%). First and second-year osteopathic medical students hold similar views on the influence of mentors and the importance of early exposure.

### Preclinical Experience in and Knowledge of Urology

The most intriguing aspect of urology selected by 47.33% of participants was surgical interventions whereas 27.33% of participants believe diagnostic procedures are the most intriguing. Sixty eight percent of respondents lack familiarity with the field of urology. Among those contemplating urology, 65.33% express a similar lack of familiarity. Upon examination of class differences, 70.37% of M1 students demonstrate unfamiliarity with urology, slightly higher than the 59.02% observed among M2 students. Notably, male, and female respondents exhibit comparable levels of unfamiliarity. When evaluating awareness of urology’s scope of practice, 65% of respondents admit to being unfamiliar with it. Among those considering urology, 57.33% share this lack of awareness regarding urology’s scope of practice.

When assessing for urology exposure preceding medical school, a substantial 76% of respondents report no exposure before entering medical school. Among females, a higher proportion (80.21%) lacks prior exposure to urology compared to males (67.92%). Furthermore, when evaluating discussions or interactions with urologists/urology residents during the preclinical year, a greater percentage of females (81.25%) indicates no such interactions compared to males (79.25%). Specifically, among those contemplating urology, a notable 84% report no discussions or interactions with urologists or urology residents during their preclinical years.

### Preferences and Attitudes Toward Preclinical Urology Exposure

A significantly greater percentage of preclinical students (56.52%) are considering urology compared to clinical students who are actively on rotations (28.57%) (*p* = 0.0064). A greater percentage of males (64.15%) are considering urology compared to females (42.71%) (*p*= 0.0164). Although not statistically significant, a greater percentage of M1 (62.96%) are considering urology compared to M2 (50.82%). 89% of respondents believe that early urology exposure is useful in deciding whether to pursue urology as a specialty. Furthermore, of the students that are considering urology, 98.67% believe that early urology exposure is useful. This was significantly greater than that of respondents not considering urology (78.67%) (*p =* 0.0001). (Figure 1)

**Figure 1.**
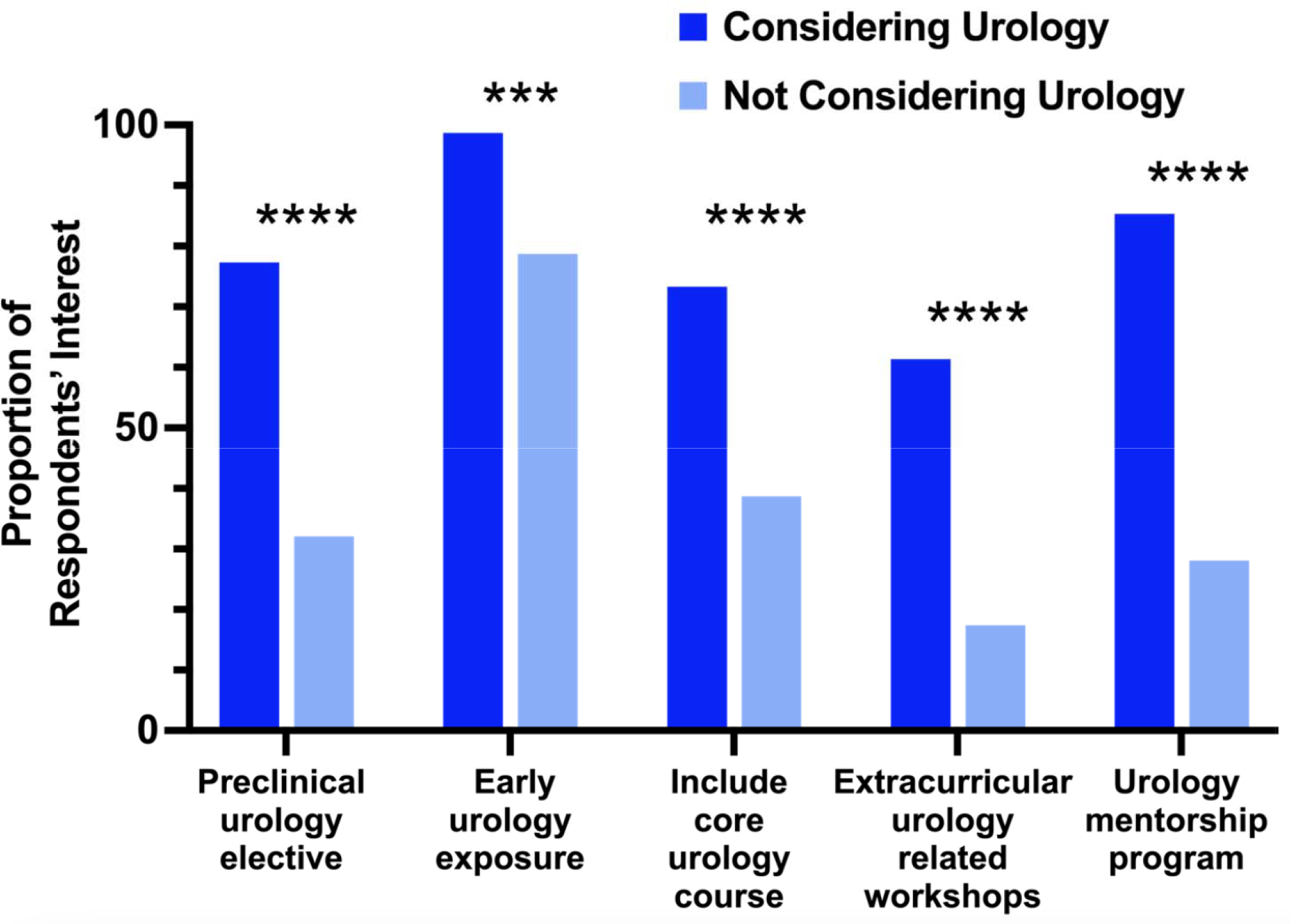
Proportion of preclinical urology exposure among respondents considering urology. Considering urology, n=75; Not considering urology, n=75. ^***^p=0.0001; ^****^p<0.0001.

In evaluating the inclination to pursue a preclinical urology elective if offered the opportunity, individuals considering urology (77%) exhibit significantly greater interest compared to those not considering urology (32%) (*p* < 0.0001). (Figure 1) Similar responses were observed between male and female groups, as well as among M1 and M2 participants. When asked whether or not to include a core urology course as part of the preclinical curriculum, 56% would include one. No differences were observed in male and female responses. However, a significantly greater percentage of M1 (68.52%) would include a core urology course compared to M2 (49.18%) (*p* = 0.0395). Moreover, of those that are considering urology, 73.33% believe that a core urology course should be included contrary to only 38.67% of those not considering urology share the same view (*p* < 0.0001). (Figure 1)

### Interest in Urology-Related Extracurricular Activities and Mentorship Programs

When looking at osteopathic medical student’s interest in attending extracurricular urology-related workshops, seminars, or conferences during preclinical years, those considering urology (61.33%) were significantly more interested compared to those not considering urology (17.33%) (*p* < 0.0001). (Figure 1) There were no differences in interest in male vs. female respondents, students currently in preclinical years vs. those in clinical years, and between first and second year osteopathic medical students. Respondents’ interest in participation in a urology mentorship program where preclinical osteopathic medical students are paired with urologists was also assessed. Those that are considering urology (85.33%) had significantly higher interest compared to those that were not considering urology (28%) (*p* < 0.0001). (Figure 1) There were no significant differences observed in the responses between the other three comparison groups.

Opinions regarding the most important step to enhance exposure and interest in urology during preclinical years were variable. Of the respondents that are considering urology, 24% believe that offering elective rotations were the most important and 24% believe that mentorship programs with urologists were the most important. Twenty percent believed that organizing urology-related workshops or simulations was the most important.

## DISCUSSION

The current study assessed osteopathic medical students’ perspectives on preclinical urology exposure and interest in approaches to increase exposure. The findings indicate that personal interest and passion, as well as lifestyle and work-life balance, are considered to be of the greatest importance when choosing a medical specialty. This result aligns with findings from studies in the literature. One study found that specialty appeal was the most chosen factor, and family time with fewer on-call duties was also important.^11^ Another study found that job satisfaction and “lifestyle following training” were the highest-rated considerations when choosing a specialty.^12^ While the present findings support previous results on overall medical specialty choice, the present study expand the scope by further investigating urology-specific perspectives.

Upon assessment of respondents’ familiarity with and consideration of urology, a greater percentage of first year students lacked familiarity compared to second year students. This discrepancy may be attributed to the fact that first-year students have not yet covered the material in their curriculum, considering that this survey was disseminated during their initial semester of medical school. Interestingly, a study found that medical students in their first month of school, have already contemplated a medical specialty preference despite minimal exposure.^12^ However, these students were open to considering other specialties. Concurrently, the present findings reveal that a lower proportion of osteopathic medical students currently in their clinical years are considering urology compared to those in the preclinical years. This result underscores the significance of early exposure to urology during the preclinical phase.

Of the respondents that are considering urology, more than half lacked awareness of urology’s scope and 84% had no preclinical discussions with urologists. This result highlights the need to increase preclinical urology exposure. Studies have corroborated this finding by indicating that urology interest groups, research opportunities between first and second year, and mentorship are strategies to encourage medical students’ interest in urology.^8^ While urology is heavily research driven field, a study found that osteopathic medical student face barriers to attaining research experiences that can make them less competitive and also affect the specialty that they ultimately want to end up in.^13^ The present results indicate that of those considering urology, inclusion of preclinical urology elective, core urology course, extracurricular urology related workshops, and a urology mentorship program is of high interest. These results may provide a basis for designing the preclinical medical education curriculum, aiming to enhance urology exposure among osteopathic medical students. This initiative may contribute to reducing the disparity in urology match rates between osteopathic and allopathic medical students.

When examining the gender cohorts, the data reveal similarities in the responses between male and female participants. Notably, a statistically significant higher proportion of males express an inclination towards urology compared to their female counterparts. Though urology is one of the most male dominated subspecialties, females exhibit a considerable interest in this specialty and gender disparities currently exist. Among practicing female surgeons, urology had one of the lowest representations and the growth rate for entering female urology residents falls behind compared to most specialties.^14^ A study found that females are mor**e** interested in pursuing OB/GYN and pediatrics than surgery.^15^ According to the 2021 AUA census data, females represent 10.9% of practicing urologists in the United States with 8.1% in the southeastern section. The present study contributes to the existing literature by emphasizing that there are inherent barriers for female individuals pursuing urology and targeting this at the medical school level is crucial. It is important to note that our study only included one respondent identifying as “Other”, preventing us from discerning perspectives from non-binary gender-identifying individuals. Further investigation may be warranted to better understand potential barriers in medical education for this demographic.

There are several limitations inherent to the present study. Firstly, the investigation exclusively captured the perspectives of students from a single osteopathic medical institution located in Florida. Considering that there are currently 37 osteopathic medical schools in the United States and only two in Florida, there may be distinct institutional experiences that could yield contrasting results compared to this study. Additionally, regional variations, particularly in the number of practicing urologists, could contribute to differences in the level of exposure students receive at their respective institutions. Based on the 2021 AUA census data, there were 13,790 number of urologists practicing in the United States, with 81.9% in the northeastern section and 59.5% in the southeastern section. Therefore, the level of exposure reported in our study might be influenced by these regional disparities. Another limitation is that a majority of the respondents were preclinical students which may not entirely encompass the perspectives of those in their clinical years. However, given that the survey questionnaire was designed to ascertain experiences specific to past or current experiences during the preclinical years, this is adequate to gain insights into perspectives on urology exposure during this particular phase of their education.

An avenue for future research involves exploring the perspectives of osteopathic medical students regarding preclinical urology exposure on a longitudinal scale. Such a study could shed light on how individual viewpoints evolve as students transition into their clinical rotations and how their exposure to urology in the preclinical years might influence their choice of specialty during the application process.

## CONCLUSION

Currently, there is a disparity in the representation of osteopathic and allopathic urologists. Insight into the perspectives of osteopathic medical students’ urology preclinical exposure is important for identifying barriers to pursuing urology and strategies to enhance urology exposure prior to clinical rotations. Results show that there is a lack of familiarity and knowledge of urology among osteopathic medical students considering urology in the preclinical years and early preclinical exposure to urology is important. Increased efforts should be made at the preclinical curriculum level to engage female individuals in urology exposure. Targeted early preclinical educational strategies are also needed to provide access for heightened urology mentorship and urology research opportunities for osteopathic medical students. Additional longitudinal research is needed to determine the sustained impact of early preclinical urology exposure on clinical rotation elective choices and residency match outcomes of osteopathic medical students.

**Supplementary Table S1:**
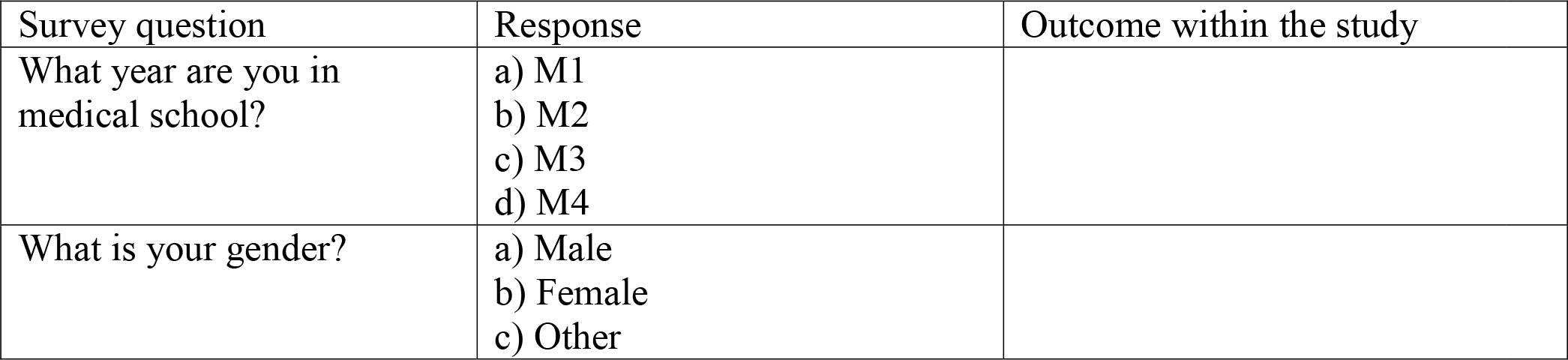
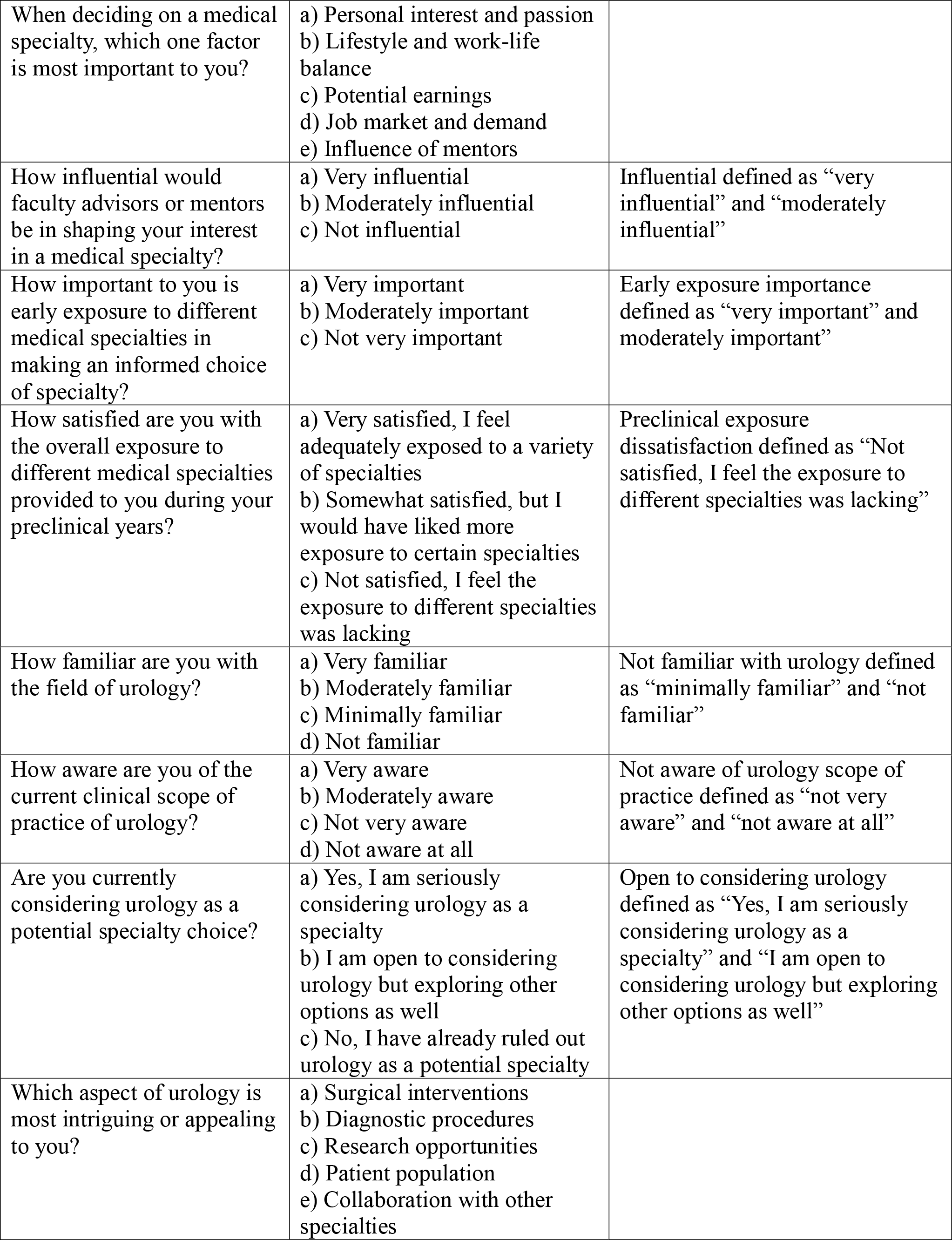
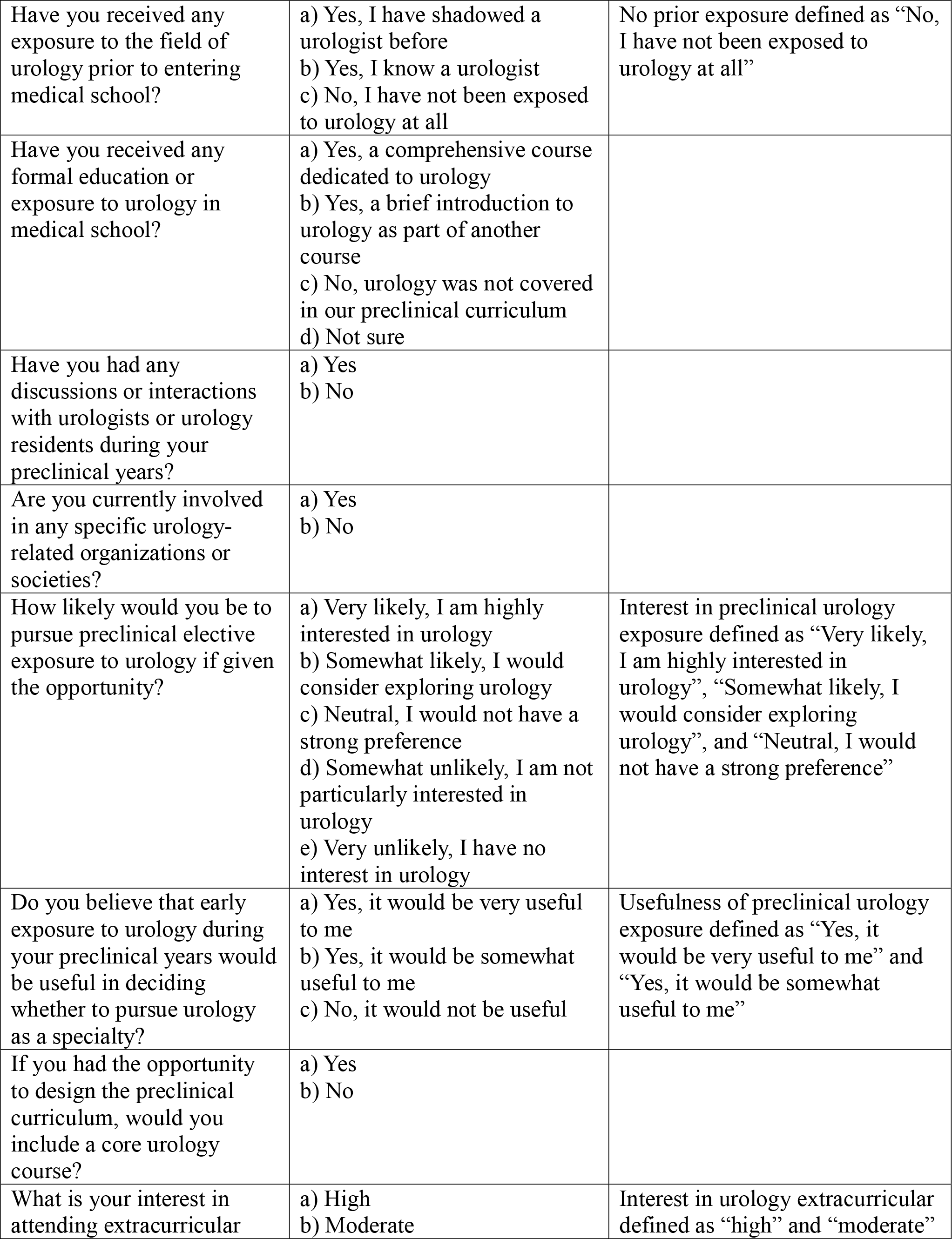
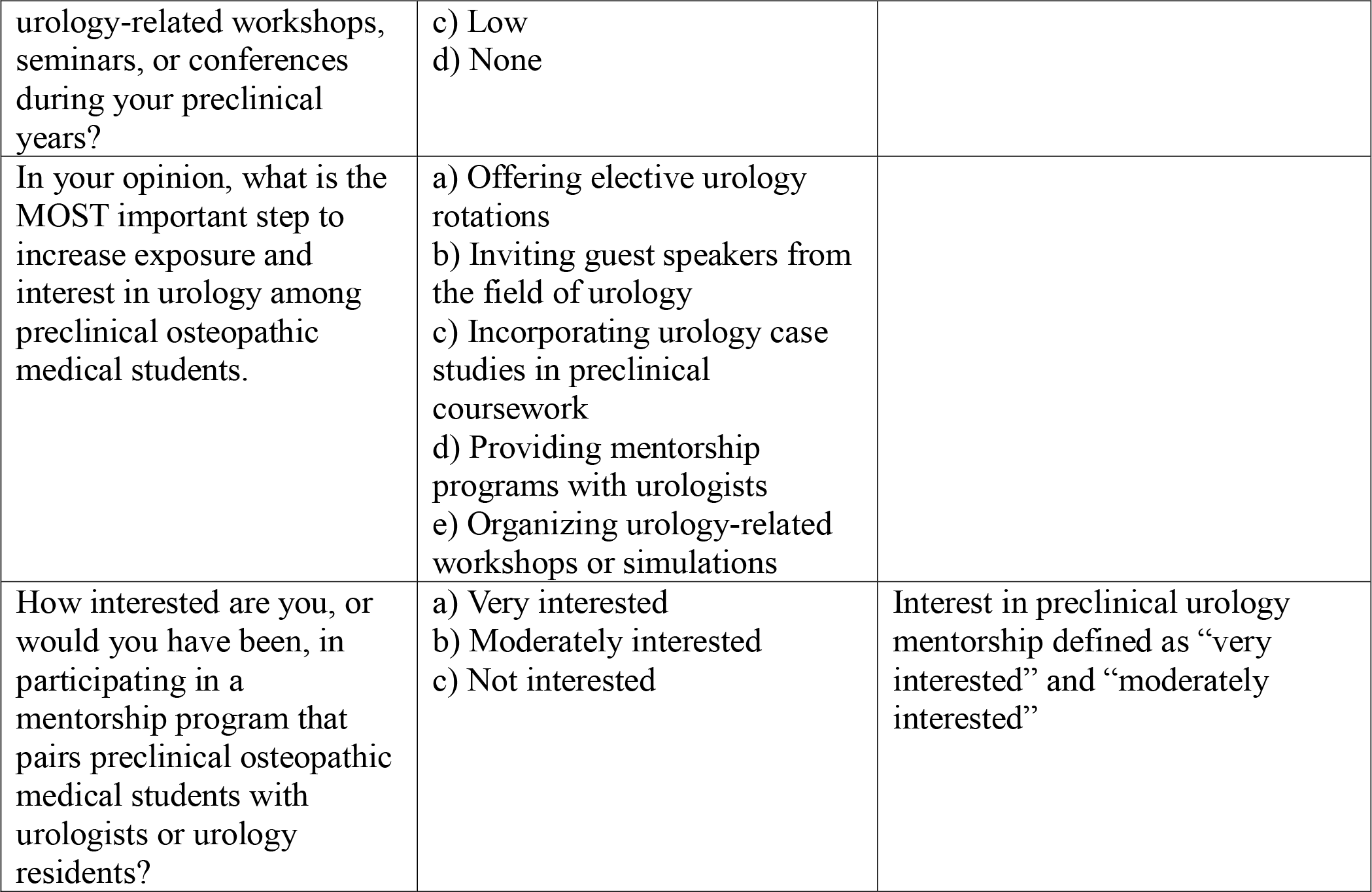
Participant survey questionnaire and schema of outcome ascertainment

## Data Availability

All data produced in the present work are contained in the manuscript

## Ethical Approval

This study was determined to be exempt from IRB review (NSU IRB Study: 2023-39)

## Informed Consent

Informed consent was not obtained.

## Author Contributions

RW and HNM provided substantial contributions to conception and design, acquisition of data, or analysis and interpretation of data; RW and HNM drafted the article or revised it critically for important intellectual content; RW and HNM gave final approval of the version of the article to be published; and RW and HNM agree to be accountable for all aspects of the work in ensuring that questions related to the accuracy or integrity of any part of the work are appropriately investigated and resolved.

